# Monoclonal Antibody Treatment of Breakthrough COVID-19 in Fully Vaccinated Individuals with High-Risk Comorbidities

**DOI:** 10.1101/2021.10.19.21265222

**Authors:** Dennis M. Bierle, Ravindra Ganesh, Sidna Tulledge-Scheitel, Sara N. Hanson, Lori L. Arndt, Caroline G. Wilker, Raymund R. Razonable

**Affiliations:** Mayo Clinic, Rochester, MN; Mayo Clinic Health Systems, Mankato, MN; Mayo Clinic Health Systems, Eau Claire, WI; Mayo Clinic Health Systems – Franciscan Healthcare, LaCrosse, WI

**Keywords:** breakthrough covid-19, casirivimab-imdevimab, vaccination, covid-19, SARS-CoV-2, outcome, hospitalization, casirivimab, imdevimab, outcomes

## Abstract

Breakthrough COVID-19 may occur in fully vaccinated persons. In this cohort of 1395 persons (mean age, 54.3 years; 60% female; median body mass index, 30.7) who developed breakthrough COVID-19, there were 107 (7.7%) who required hospitalization by day 28. Hospitalization was significantly associated with the number of medical comorbidities. Anti-spike monoclonal antibody treatment was significantly associated with a lower risk of hospitalization (Odds Ratio: 0.227; 95% confidence interval, 0.128 - 0.403; p<0.001). The number needed to treat to prevent one hospitalization was 225 among the lowest-risk patient group compared to 4 among the groups with highest numbers of medical comorbidity.

**summary:** Breakthrough COVID-19 may occur among vaccinated individuals with a high number of medical comorbidities, especially during the surge of SARS-CoV-2 Delta variant. Treatment with anti-spike monoclonal antibody was associated with significantly lower rates of hospitalization and oxygen supplementation.

## Introduction

COVID-19 vaccination is the primary strategy to reduce severe acute respiratory syndrome coronavirus 2 (SARS-CoV-2) outbreaks and its great burden to healthcare systems ^1^. Clinical trial and real-world experience suggest high efficacy of SARS-CoV-2 vaccine for the prevention of COVID-19 ^1 2,3^. Despite the effectiveness of COVID-19 vaccines, breakthrough infections have been reported ^4^. Among healthcare workers with breakthrough COVID-19, the majority of infections were mild, although 19% had persistent symptoms ^4^. With the surge in SARS-CoV-2 B.1.617.2 (Delta) variant, there have been increasing reports of breakthrough infections among vaccinated persons.

Real-world clinical data are needed to characterize breakthrough COVID-19 in fully vaccinated persons and assess their risk of hospitalization. Moreover, there is no data as to whether vaccinated individuals with breakthrough COVID-19 would benefit from passive immunotherapy with casirivimab-imdevimab or bamlanivimab-etesevimab. In this study, we assessed the clinical outcomes of 1395 fully vaccinated adult patients with breakthrough COVID-19. We specifically investigated the rates of severe disease, as measured by oxygen requirement and need for hospitalization, and we compared these outcomes between patients who received or did not receive treatment with anti-spike monoclonal antibody.

## Methods

### Patient Population and Study Design

After approval by the Mayo Clinic Institutional Review Board, this retrospective study enrolled fully vaccinated patients with breakthrough COVID-19 who were screened for eligibility for treatment with bamlanivimab (Eli Lilly, Indianapolis, IN), bamlanivimab-etesevimab (Eli Lilly, Indianapolis, IN) or casirivimab-imdevimab (Regeneron, New York) as single infusion from January to August 16, 2021. All patients developed PCR-confirmed breakthrough COVID-19 at least 2 weeks after the second of two doses of SARS-CoV-2 mRNA vaccine (Pfizer-BioNTech or Moderna) or the single dose of Ad26.COV2.S (Johnson and Johnson) vaccine.

All patients were identified after being screened by the Mayo Clinic Monoclonal Antibody Treatment Program in Mayo Clinic sites in the Midwest, as previously described.^5^ All patients had at least 28 days of follow up after the COVID-19 diagnosis. Only patients who provided authorization for the use of their medical records for research purposes were included. The conduct of this study was in compliance with the aims of Strengthening the Reporting of Observational Studies in Epidemiology (STROBE)^6^.

### Clinical Variables

The demographic and clinical characteristics of all patients, including COVID-19 vaccination status and date of vaccination, were collected by a review of their Electronic Health Records. Medical comorbidities and other characteristics that portend high-risk for progression to severe COVID-19 were recorded. The degree of medical comorbidity was assessed using Monoclonal Antibody Screening Score (MASS), a clinical criteria for risk of severe outcomes that has been previously described ^7,8^. MASS assigned points to characteristics and comorbidity that made a person eligible for monoclonal antibody, according to the initial Food and Drug Administration (FDA) Emergency Use Authorization (EUA) guidance issued in November 2020 ^9^. In May 2021, the US FDA expanded the use of monoclonal antibodies to include lower-risk persons such as those with body mass index of 25 kg/m^2^. For the purposes of this study, MASS was used as a measure of medical comorbidity burden, as previously described ^7,8^.

### Clinical Outcomes

The primary outcome for this study was the rate of all-cause hospitalization by day 28 after the diagnosis of breakthrough COVID-19. The need for oxygen supplementation was recorded as an indicator of severe disease. All 1395 patients had completed at least 28 days of follow up by the end of this study on September 15, 2021.

### Statistical Analysis

Data are presented in aggregate using descriptive statistics. The differences in the outcomes between two subgroups of patients, defined based on receipt or non-receipt of monoclonal antibodies, were compared using descriptive statistics. To calculate the proportion of patients who experienced 28-day hospitalization or use of oxygen supplementation, each patient who experienced one of the two outcomes was assigned to the numerator with the total population as the denominator. Simple proportions were calculated by receipt of monoclonal antibody therapy and stratified by MASS score. Differences in these proportions were assessed using Chi Square test for independence for categorical variables or a t-test for continuous variables after ensuring testing parameters were met. Fisher Exact test was used if testing assumptions were not achieved. Crude odds ratios (OR) with 95% confidence intervals (CIs), absolute risk reduction and number needed to treat were calculated to compare associations of patient characteristics with outcomes; P < 0.05 was deemed statistically significant. The association between monoclonal antibody therapy and hospitalization within 28 days was assessed with logistic regression with adjustment for MASS. Data was analyzed using Microsoft Excel (Microsoft Excel for Office 365, Version 16.0.13127.21766, Microsoft Corporation, Redmond, WA) and BlueSky Statistics (Commercial Server Edition, Version 7.40).

## Results

The total population consisted of 1395 fully vaccinated patients (mean age, 54.3 years; 39.6% male) with breakthrough COVID-19 and were screened for monoclonal antibody therapy, under US FDA EUA guidance. The majority of breakthrough SARS-CoV-2 infections (n=1002; 71.8%) occurred during a 6-week period from July 1 to August 16, 2021, when the Delta variant was predominant in our region. The most common medical comorbidities were hypertension (34.5%), cardiovascular disease (13.8%), chronic pulmonary disease (13.6%), chronic kidney disease (10.2%), and cancer (10.7%) (**Table 1**). Most patients (69.8%) received the Pfizer-BioNTech SARS-CoV-2 mRNA vaccine.

**Table 1.** Demographic and Clinical Characteristics of 1395 Vaccinated Persons with Breakthrough Coronavirus Disease-2019, Mayo Clinic in the Midwest, January 1 – August 16, 2021

| Characteristics | Number of patients (percentage), unless otherwise specified |
| --- | --- |
| Age, in years, mean ± standard deviation | 54.3 ± 18.5 |
| Male sex | 553 (39.64%) |
| Body mass index, mean ± standard deviation | 30.7 ± 8.8 |
| Cancer | 149 (10.68%) |
| Cardiac disease | 192 (13.76%) |
| Chronic kidney disease | 142 (10.18%) |
| Diabetes mellitus | 208 (14.91%) |
| Hypertension | 481 (34.48%) |
| Pulmonary disease | 190 (13.62%) |
| Covid-19 vaccine type |  |
| Ad26.COV2.S vaccine (Johnson and Johnson) | 133 (9.53%) |
| BNT162b2 vaccine (Pfizer-BioNTech) | 974 (69.82%) |
| mRNA-1273 vaccine (Moderna) | 288 (20.65%) |
| Time between full vaccination date and breakthrough COVID-19, in days; mean ± standard deviation | 120 ± 53.7 |
| Monoclonal antibody treatment |  |
| Bamlanivimab or bamlanivimab-etesevimab | 43 (3.08%) |
| Casirivimab-imdevimab | 484 (34.70%) |
| No treatment | 868 (62.22%) |

Breakthrough COVID-19 occurred at mean of 120 days after full completion of the vaccination series. At 28 days after breakthrough COVID-19 diagnosis, 107 patients (7.7%) had progressed to require hospitalization. The rate of hospitalization was significantly higher among patients with higher medical comorbidity score (**Table 2** and **Supplementary Table**).

**Table 2.** Differences in Rates of Hospitalization among 1395 Vaccinated Persons with Breakthrough Coronavirus Disease-2019 According to Treatment with Anti-spike Monoclonal Antibody

| MASS* | Monoclonal antibody treatment |  | No treatment |  | Relative Risk | Odds Ratio (95% CI) | Absolute Risk Reduction | NNT |
| --- | --- | --- | --- | --- | --- | --- | --- | --- |
|  | Number (hospitalized) | Rate | Number (hospitalized) | Rate |  |  |  |  |
| 0 | 117 (2) | 0.017 | 428 (9) | 0.021 | 0.813 | 0.809 (0.173-3.800) | 0.004 | 255 |
| 1 | 83 (0) | 0 | 86 (2) | 0.023 | 0 | 0.202 (0.010-4.280) | 0.023 | 43 |
| 2 | 49 (1) | 0.020 | 60 (4) | 0.067 | 0.306 | 0.292 (0.032-2.700) | 0.046 | 22 |
| 3 | 92 (3) | 0.032 | 57 (7) | 0.123 | 0.266 | 0.241 (0.060-0.973) | 0.090 | 12 |
| 4 | 39 (0) | 0 | 53 (8) | 0.151 | 0 | 0.068 (0.125-0.028) | 0.151 | 7 |
| ≥4 (pooled) | 186 (8) | 0.043 | 237 (71) | 0.300 | 0.143 | 0.105 (0.049-0.225) | 0.257 | 4 |
| 5 | 41 (2) | 0.048 | 72 (21) | 0.292 | 0.167 | 0.125 (0.243-0.040) | 0.243 | 5 |
| 6 | 37 (2) | 0.054 | 21 (4) | 0.190 | 0.284 | 0.243 (0.152-0.031) | 0.136 | 8 |
| 7 | 26 (2) | 0.076 | 34 (12) | 0.353 | 0.218 | 0.153 (0.058-0.128) | 0.276 | 4 |
| ≥8 (pooled) | 43 (2) | 0.046 | 57 (26) | 0.456 | 0.102 | 0.058 (0.013-0.264) | 0.409 | 3 |

\*MASS assigned a score to each of the FDA EUA eligibility criteria (November 2020), as follows: age ≥65 (2 points), BMI ≥35 (1 point), diabetes mellitus (2 points), chronic kidney disease (3 points), cardiovascular disease in a patient ≥55 years (2 points), chronic respiratory disease in a patient ≥55 years (2 points), hypertension in a patient ≥55 years (1 point), and immunocompromised status (3 points).

**Table 2** stratifies the population into those who received (n=527; 37.8%) versus those who did not receive (n=868; 62.2%) treatment with anti-spike monoclonal antibody. Among those who received treatment, the majority were treated with casirivimab-imdevimab, including all patients diagnosed after May 7, 2021. Patients received the monoclonal antibody treatment at a median of 5 days (interquartile range, 3-6 days) from symptom onset and 2 days (interquartile range, 1-3 days) from testing. Overall, the rate of hospitalization was 2.65% among patients treated with monoclonal antibody compared to 10.7% among those who did not receive therapy (Odds Ratio [OR]: 0.227; 95% confidence [CI] interval, 0.128 - 0.403; p<0.001). Controlling for MASS, monoclonal antibody therapy was associated with a lower rate of hospitalization (OR: 0.14; 95% CI, 0.079-0.265; p<0.0001). The number needed to treat (NNT) to prevent one hospitalization correlated with the degree of medical comorbidity. NNT was 255 among vaccinated persons without high-risk medical comorbidity while NNT was between 3 and 8 among patients with multiple high-risk medical conditions (**Table** 3, polled NNT of 4 for patient groups with MASS ≥4).

Sixty-one patients (4.4%) developed hypoxia and required oxygen supplementation. Anti-spike monoclonal antibody treatment was significantly associated with lower rates of hypoxia when compared to those who did not received monoclonal antibody (0.95% versus 6.45%; OR, 0.139; 95% CI, 0.055 – 0.349; p <0.001). The majority (n=56; 91.8%) of patients who required oxygen supplementation did not receive monoclonal antibody treatment. Five (0.36%) patients required admission to the intensive care unit (ICU), none of whom were treated with monoclonal antibody. No death was reported among 1395 vaccinated patients with breakthrough COVID-19.

## Discussion

This study provides several observations that are relevant in the current era of the SARS-CoV-2 pandemic that is dominated by the Delta variant causing breakthrough COVID-19 in fully vaccinated individuals. First, this study observed that, among patients with breakthrough COVID-19, 8% developed clinical progression to require hospitalization, including some needing oxygen supplementation. This finding emphasizes the need for other public health strategies (e.g., use of face mask and avoidance of large gatherings) for all individuals, even among vaccinated individuals, during periods characterized by high community SARS-CoV-2 Delta transmission.

Second, this study demonstrates that the risk of hospitalization among breakthrough COVID-19 cases varies according to the degree of medical comorbidity. In our cohort, obesity, hypertension, diabetes, cancer, chronic lung, kidney and cardiac conditions accounted for majority of comorbidities. This study demonstrates that individuals who possessed multiple comorbidities, as measured by MASS, were at higher risk of hospitalization. Some of the multi-comorbidity patients in this study included those with cancer and immune compromised patients who may have suboptimal response to COVID-19 vaccine. This finding supports the current FDA EUA for post exposure prophylaxis in high risk patients who have been fully immunized but are at high risk of not mounting a robust immune response secondary to immunocompromised state.

Third, this study provides clinical evidence of the benefit of augmented passive immunotherapy among fully vaccinated patients with breakthrough COVID-19. This benefit is especially evident among those with a high number of medical comorbidities. As indicated above, it is possible that these high-risk individuals may have had suboptimal response to COVID-19 vaccine. Supplementing these patients with monoclonal antibody immunotherapy may have mitigated their risk of clinical disease progression ^10^. Monoclonal antibody treatment of breakthrough COVID-19 resulted in a lower risk of hospitalization, and a reduced need for oxygen supplementation among hospitalized patients. This observation is consistent with our previous report that associated monoclonal antibody treatment with reduced rates of hospitalization compared to those who did not receive treatment ^11,12^. The vast majority of our cohort developed breakthrough COVID-19 during the Delta surge, and this finding provides the clinical correlate to experimental data that suggested that casirivimab-imdevimab combination (which was used solely after May 7, 2021) retains efficacy against SARS-CoV-2 B.1.617.2, which is assumed as main cause of the breakthrough cases^13,14^.

This was a retrospective study with the inherent limitations to this study design. For example, we would not have captured treated patients who may have been hospitalized at outside institutions. However, we believe that this is not a considerable number since our program enrolls high-risk patients to a remote monitoring program, thereby ensuring continued contact with these patients until symptom resolution^5^. Our population was predominantly Caucasian, and our findings may not be generalizable to other patient populations. Most patients received treatment with casirivimab-imdevimab, limiting generalizability to other monoclonal antibodies such as bamlanivimab-etesevimab and sotrovimab. The study observations should also be interpreted in the context that it was performed in patients screened and treated in a single healthcare system. These limitations are counterbalanced by having a relatively large cohort of breakthrough COVID-19 cases identified in our healthcare system, with many identified during the period of the Delta surge. This large patient cohort allowed for robust statistical analysis of the potential efficacy of treatment with monoclonal antibody therapy.

## Conclusion

Fully vaccinated persons, particularly those with multiple medical comorbidities, may develop breakthrough COVID-19, especially during the surge of a more communicable Delta variant. Some patients progressed to severe COVID-19 that required hospitalization, including the need of oxygen supplementation and ICU care. The severity of breakthrough COVID-19 significantly correlated with a high number of medical comorbidities. Reassuringly, no death was reported. Early treatment with anti-spike monoclonal antibody was associated with a significantly lower rate of severe COVID-19 that would require hospitalization.

## Data Availability

All data produced in the present study are available upon reasonable request to the authors

## Footnotes

### Ethical approval

This study was approved by the Mayo Clinic Institutional Review Board.

## Acknowledgments

The author would like to thank all members of the Mayo Clinic Monoclonal Antibody Treatment Program.

---

**Supplementary Table.** Rates of Hospitalization among 1395 Vaccinated Individuals with Breakthrough Coronavirus Disease-2019, Stratified Based on Monoclonal Antibody Screening Score

| Monoclonal Antibody Screening Score (MASS) | Number of Patients | Number of Hospitalized Patients | Odds Ratio (95% Confidence Interval) | P value |
| --- | --- | --- | --- | --- |
| 0 | 545 | 11 | 1 | - |
| 1 | 169 | 2 | 0.58 (0.13-2.65) | 0.478 |
| 2 | 109 | 5 | 2.33 (0.79-6.86) | 0.113 |
| 3 | 149 | 10 | 3.49 (1.45-8.39) | 0.003 |
| 4 | 92 | 8 | 4.6 (1.81-11.83) | < 0.001 |
| 5 | 113 | 23 | 12.41 (5.85-26.33) | < 0.001 |
| 6 | 58 | 6 | 5.60 (1.99-15.76) | < 0.001 |
| 7 | 60 | 14 | 14.77 (6.35-34.40) | < 0.001 |
| 8 | 41 | 9 | 13.65 (5.28-35.32) | < 0.001 |
| 9 | 17 | 8 | 43.15 (14.02-132.79) | < 0.001 |
| 10 | 25 | 7 | 18.88 (6.56-54.36) | < 0.001 |
| 11 | 11 | 3 | 18.20 (4.25-78.00) | < 0.001 |
| 12 | 4 | 1 | 16.18 (1.55-168.09) | < 0.001 |
| 13 | 2 | 0 | 0 | 0.8392 |
| Total | 1395 | 107 |  |  |

MASS assigned a score to each of the FDA EUA eligibility criteria (November 2020), as follows: age $\geq 65$ (2 points), BMI $\geq 35$ (1 point), diabetes mellitus (2 points), chronic kidney disease (3 points), cardiovascular disease in a patient $\geq 55$ years (2 points), chronic respiratory disease in a patient $\geq 55$ years (2 points), hypertension in a patient $\geq 55$ years (1 point), and immunocompromised status (3 points).

